# Changes in serum TSH across the adult lifespan: 22-year follow-up of the HUNT Study

**DOI:** 10.64898/2025.12.14.25342222

**Authors:** Bjørn O. Åsvold, Marion Denos, Peter N. Taylor, Salman Razvi, Trine Bjøro, Ben M. Brumpton, Eirin B. Haug

**Author notes:** Corresponding author: Bjørn Olav Åsvold, MD PhD, HUNT Center for Molecular and Clinical Epidemiology, Department of Public Health and Nursing, NTNU, Norwegian University of Science and Technology, Postboks 8905 MTFS, NO-7491 Trondheim Norway.

## Abstract

**Objective:** Some evidence suggests that higher serum TSH may be a part of normal aging, but current studies are limited to 13-year follow-up. We examined TSH changes during 22 years of follow-up across the adult lifespan.

**Design:** Longitudinal analyses of the population-based HUNT Study in Norway, with TSH measurements from 1995-97, 2006-08 and 2017-19.

**Methods:** In the overall population and in individuals without thyroid medication or disease, we estimated i) geometric mean serum TSH by age, integrating cross-sectional and longitudinal measurements using linear mixed models, ii) percentiles of the TSH distribution by age, and iii) within-individual TSH change during follow-up, expressed by geometric mean ratios (GMR) reflecting the fold change in geometric mean TSH.

**Results:** We included 136,925 TSH measurements among 84,342 participants, of whom 40,615 had ≥2 measurements and 13,613 had ∼22-year follow-up. Mean TSH was higher at older age in men, but weaker and less consistent in women. The TSH distribution widened at older age in men and women. Among individuals without thyroid medication or disease, mean TSH increased modestly by 0.13 mIU/L (GMR 1.09; 95%CI 1.08,1.11) during 22-year follow-up in men, but not in women (GMR 0.99; 95%CI 0.98,1.00). This increase was stronger at 0.5 mIU/L in men aged ≥70 years at baseline (GMR 1.32; 95%CI 1.18,1.48).

**Conclusions:** Mean serum TSH increased with age in older men, but showed only modest or no age-related change in younger men and in women. The wider TSH distribution at older age supports the need for age-specific TSH reference ranges.

**Significance statement:** Previous evidence of higher TSH concentrations at older age comes from cross-sectional studies and longitudinal studies with up to 13-year follow-up. We utilized a large population-based study to extend the evidence on within-individual TSH changes to ∼22-year follow-up across the adult lifespan. Mean TSH increased with age in men, modestly at 0.13 mIU/L overall, but stronger at 0.5 mIU/L in men followed from their 70s to their 90s. In women, mean serum TSH appeared stable during follow-up, but more frequent thyroid hormone supplementation may have skewed the TSH distribution away from higher, but still physiological levels. The TSH distribution widened at older age in both men and women, supporting the need for age-specific TSH reference ranges.

## Introduction

In several cross-sectional population-based studies, older age has been associated with higher mean serum TSH concentrations, even among individuals without evidence of thyroid disease.^1–4^ These observations suggest that higher TSH concentrations, indicating a higher TSH setpoint, may be physiologically normal at older age.^5^ However, such cross-sectional associations may be vulnerable to selection bias, and could be due to effects of birth cohort rather than age (e.g. if birth cohorts differ in their life course history of iodine status). In contrast, longitudinal studies can more validly assess within-individual age-related changes, but have yielded conflicting results.^6^ Some studies of adult^7^ or elderly^8,9^ individuals observed increases in TSH concentrations during 8-13 years of follow-up, whereas other studies of 5-7 years of follow-up did not observe changes in TSH.^10–12^

The population-based Trøndelag Health Study (the HUNT Study) in Norway is one of the studies in which older age has cross-sectionally been associated with higher TSH concentrations.^4,13^ The HUNT study cohort has subsequently been followed with serum TSH measurements during more than 20 years. Utilizing this longitudinal population-based cohort, we aimed to estimate age-related changes in TSH concentrations.

## Materials and methods

### Study population

The HUNT Study is a series of population-based health surveys carried out in the Nord-Trøndelag area in Norway.^14^ In each survey, all current residents aged ≥20 years were invited to participate, and the study thus provides a combination of longitudinal (among people participating in more than one survey) and cross-sectional (among people participating in one survey only) data. The current study includes participants from the HUNT2 (1995-97), HUNT3 (2006-08) and HUNT4 (2017-19) surveys, which included measurement of thyroid function. The participation rate was 69% (n=65,228) in HUNT2, 54% (n=50,800) in HUNT3, and 54% (n=56,042) in HUNT4. Dairy products and lean fish are the main sources of iodine in Norway.^15^ The population has generally been considered to be iodine sufficient,^16^ but analyses of HUNT4 suggested borderline iodine deficiency with median urinary iodine concentration of 97 µg/L. The median estimated iodine intake at HUNT4 was 171 µg/day overall, 179 µg/day in men and 165 µg/day in women.^17^

In HUNT2, serum TSH was measured in subsamples of the population, including all women aged ≥40 years, a 50% random sample of men aged ≥40 years, and 5% random samples of women and men <40 years of age. In total, TSH concentrations were measured in 33,945 participants from these samples. Among them, we excluded 31 who did not reply to the questionnaire items on thyroid diseases, leaving 33,914 participants for analysis. In HUNT3 and HUNT4, TSH measurement was planned for all participants, and TSH concentrations were measured in 49,175 HUNT3 participants and 53,836 HUNT4 participants, who were included in the present analyses.

### Thyroid function measurements

In HUNT2, individuals with serum TSH >4.0 mIU/L also had their serum free thyroxine and TPO antibody concentrations measured, and serum free thyroxine was also measured if serum TSH was <0.20 mIU/L. The measurements were performed at the Hormone Laboratory at Oslo University Hospital, using DELFIA hTSH Ultra and DELFIA FT4 from Wallac Oy (Turku, Finland) and a luminoimmunoassay for TPO antibodies from B.R.A.H.M.S. Diagnostica GmbH (Berlin, Germany). The reference ranges were 0.2–4.5 mIU/L for TSH, 8–20pmol/L for free thyroxine and <200 U/mL for TPO antibodies.

In HUNT3, individuals with serum TSH <0.10 mIU/L or >3.00 mIU/L had their serum free thyroxine and TPO antibody concentrations measured. TSH, free thyroxine and TPO antibody concentrations were measured at Levanger Hospital, Nord-Trøndelag Hospital Trust, using chemiluminescent microparticle immunoassays on an Architect ci8200 from Abbott, with reagents obtained from Architect iSystem (Abbott Ireland, Longford, Ireland; and Abbott Laboratories). The reference ranges were 0.20–4.5 mIU/L for TSH, 9.0-19.0 pmol/L for free thyroxine and ≤5.6 U/mL for TPO antibodies. The TSH measurement methods in HUNT2 and HUNT3 were compared using blood samples from 94 individuals. For TSH concentrations below 5mIU/l, the results did not systematically differ between the methods. For TSH concentrations above 5 mIU/L, the values were, on average, 3.7% lower when using the Architect method (HUNT3) than when using the DELFIA method (HUNT2).^18^

In HUNT4, individuals with serum TSH <0.10 mIU/L or >3.60 mIU/L had their serum free thyroxine and TPO antibody concentrations measured. TSH, free thyroxine and TPO antibody concentrations were measured at Levanger Hospital using the same methods as in HUNT3. However, different TSH reagent kits were used in HUNT3 (7K62 (68-5842/R1) and HUNT4 (2K62 TSH), and the laboratory TSH reference range at HUNT4 was 0.5-3.6 mIU/L.^13^ In HUNT3 and HUNT4 subpopulations (each of 9208 individuals) matched on sex, age, self-reported general health and month of blood sampling, median serum TSH in HUNT4 was 0.02106 + 0.8020 x serum TSH of HUNT3.^13^ To enable comparison across the HUNT surveys, we therefore corrected the TSH measurements in HUNT4 using the formula: corrected serum TSH in HUNT4 = (measured serum TSH in HUNT4 - 0.02106) / 0.8020. We apply these corrected HUNT4 values in all analyses and descriptions from here onwards. The free thyroxine measurements in HUNT4 (reference range 10-17 pmol/L) were performed in 2024 in serum samples stored at −80 °C in the HUNT Biobank.^19^

### Assessment of thyroid medication and disease

In HUNT2, participants were queried about thyroid diseases, including ever use of levothyroxine or thionamides. To obtain information about the use of thyroid medication at the time of HUNT3 and HUNT4, we used the unique 11-digit identity number of every Norwegian citizen to link the HUNT data with information from the Norwegian Prescribed Drug Registry, which includes information on virtually all prescriptions dispensed to non-institutionalized inhabitants in Norway since 2004. From this registry, we obtained individual information on dates on which thyroid hormones, thionamides and amiodarone had been dispensed from pharmacies. At each survey, we classified participants as using thyroid hormone replacement therapy or thionamides based on self-report of ever use (HUNT2), or by at least one drug dispensing during the last year recorded in the Norwegian Prescribed Drug Registry (HUNT3 and HUNT4). Among individuals without such treatment, we used the following criteria to identify untreated thyroid disease: i) serum TSH <0.10 or >10.0 mIU/L, or ii) TSH >4.5 mIU/L combined with TPO antibody concentration above the reference range, or iii) TSH >4.5 mIU/L combined with serum free thyroxine below the reference range. These criteria were selected to ensure consistency across the surveys.

### Statistical analysis

First, we used linear mixed models to estimate geometric mean TSH concentrations (with 95% confidence intervals) by age. These models integrated all cross-sectional and longitudinal TSH measurements and included random intercepts to account for repeated TSH measurements within individuals. Age was expressed by a restricted cubic spline with knots at every 10 years from 20 to 90 years of age. We analyzed men and women separately, as incident use of thyroid medication strongly differed by sex in our study population.^18^ We performed the analyses for three different situations: i) To estimate the crude overall association between age and TSH concentrations in the population, we included all participants and TSH measurements; ii) To approximate the association between age and TSH concentrations that would occur without thyroid treatment, we replaced the TSH values among treated individuals: TSH was set at 4.5 mIU/L in individuals treated with thyroid hormones who had lower TSH levels, and TSH was set at 0.10 mIU/L in individuals treated with thionamides who had higher TSH levels. This probably represents conservative estimates of the TSH abnormalities that would be present without treatment. When aiming to estimate the overall TSH changes in the population, such approximation of the untreated values is likely superior to e.g. excluding treated individuals;^20^ iii) To approximate the association between age and TSH in individuals without thyroid disease, we excluded TSH measurements performed during thyroid medication (thyroid hormones or thionamides) or untreated thyroid disease. We assumed that this approach would yield the most valid estimates of physiological differences in serum TSH concentrations by age. In sensitivity analyses, we excluded TSH measurements among current daily smokers and TSH measurements performed during pregnancy or after any dispensing of amiodarone between 2004 and the date of TSH measurement.

Second, to describe the distribution of TSH concentrations across age, we used quantile regression to estimate the 10-, 25-, 50-, 75- and 90-percentiles of the serum TSH distribution by age, using similar analytical approaches as above.

Third, to estimate the within-individual changes in serum TSH during follow-up, we performed longitudinal analyses among individuals with at least two TSH measurements. We examined the change in serum TSH during the first 11 years of follow-up (from the first TSH measurement (HUNT2 or HUNT3) to the next survey (HUNT3 or HUNT4); median (range) follow-up time 11.0 (9.3-12.8) years), and during the entire 22-year follow-up (from HUNT2 to HUNT4, (median (range) follow-up time 21.8 (20.3-23.3) years). To account for inter-individual differences in follow-up time between surveys, we standardized the follow-up time to 11 and 22 years by dividing the change in log-transformed serum TSH from baseline to follow-up by the number of 11- and 22-year units of follow-up time, respectively. The change in TSH was expressed as geometric mean ratios (GMR) that express the fold change in geometric mean TSH from baseline to follow-up examination. We estimated the TSH changes overall and by 10-year age group, separately in men and women, and for similar situations as above. Finally, we calculated the mean (SD) changes in TSH during follow-up by sex and age among participants without thyroid medication or disease at baseline or follow-up, and tested whether absolute deviations from the mean TSH change varied by age.

The data were analyzed using StataMP 19.5 for Windows (StataCorp LLC, College Station, Texas). The study was approved by the Regional Committee for Medical and Health Research Ethics (REK midt 215744), and all participants gave informed consent.

## Results

The analyses included 84,342 participants with a total of 136,925 TSH measurements (Table 1). Among them, 11,968 participants had TSH information from all three HUNT surveys (median follow-up from first to last TSH measurement 21.8 years), 1645 participants had TSH measured in the HUNT2 and HUNT4 surveys only (median follow-up 21.8 years), 27,002 had TSH measured in two adjacent surveys only (HUNT2 and HUNT3, or HUNT3 and HUNT4; median follow-up 10.7 years), and 43,727 contributed with one TSH measurement only. The age at TSH measurement ranged from 19 to 102 years (Supplementary Table 1). Among 57,346 TSH measurements in men, 1254 (2.2%) were performed among men using thyroid hormone replacement or thionamides, and additional 636 (1.1%) were performed among men with biochemical evidence of untreated thyroid disease, leaving 55,456 (96.7%) measurement among men presumably without thyroid disease. Among 79,579 TSH measurements in women, the corresponding numbers were 6571 (8.3%), 1850 (2.3%), and 71,158 (89.4%), respectively.

**Table 1.** Characteristics of the study population.

| Characteristics | HUNT2 (1995-97)<sup>a</sup> |  | HUNT3 (2006-08) |  | HUNT4 (2017-19) |  |
| --- | --- | --- | --- | --- | --- | --- |
|  | Men<br>(n=10,643) | Women<br>(n=23,271) | Men<br>(n=22,354) | Women<br>(n=26,821) | Men<br>(n=24,349) | Women<br>(n=29,487) |
| Age (years), mean (SD) | 50.3 (16.7) | 50.3 (17.4) | 53.7 (15.6) | 52.8 (16.3) | 55.1 (17.2) | 54.1 (17.7) |
| Serum TSH (mIU/L), median (IQR) | 1.5 (1.1-2.1) | 1.5 (1.0-2.2) | 1.5 (1.1-2.0) | 1.4 (1.0-1.9) | 1.5 (1.1-2.0) | 1.4 (1.0-1.9) |
| Thyroid medication,<sup>b</sup> % | 1.0 | 5.0 | 2.1 | 8.4 | 2.7 | 9.2 |
| Untreated overt hypothyroidism,<sup>c</sup> % | 0.2 | 0.7 | 0.1 | 0.1 | 0.4 | 0.5 |
| Untreated overt hyperthyroidism,<sup>d</sup> % | 0.05 | 0.2 | 0.04 | 0.1 | 0.01 | 0.1 |
| Body mass index (kg/m<sup>2</sup>), mean (SD) | 26.5 (3.5) | 26.3 (4.5) | 27.5 (3.8) | 26.9 (4.8) | 27.6 (4.2) | 26.9 (5.1) |
| Current daily smokers, % | 28.5 | 29.2 | 15.0 | 19.3 | 6.9 | 9.6 |

IQR, interquartile range; SD, standard deviation; TSH, thyroid-stimulating hormone
<sup>a</sup> Participants aged <40 years were weighted due to differences in sampling probability
<sup>b</sup> Use of thyroid hormone replacement therapy or thionamides, either ever use based on self-report (HUNT2) or dispensing during the last year based on information from the Norwegian Prescribed Drug Registry (HUNT3 and HUNT4)
<sup>c</sup> Serum TSH >4.50 mIU/L combined with serum free thyroxine below the reference range, in individuals without thyroid medication
<sup>d</sup> Serum TSH <0.10 mIU/L combined with serum free thyroxine above the reference range, in individuals without thyroid medication

When integrating all cross-sectional and longitudinal TSH measurements, we observed that in men, geometric mean serum TSH was quite stable at ∼1.4 mIU/L from ages 20 to 60 years, followed by a gradual increase to ∼1.6 mIU/L at age 80 and ∼1.8 mIU/L at age 90. The age-TSH association was similar when we excluded men with thyroid medication or disease, and slightly stronger when we instead replaced TSH values among treated individuals to approximate the TSH abnormalities that would be present without treatment (Figure 1A, Supplementary Table 2). In women, geometric mean serum TSH was quite similar across all ages in the overall study population, but increased from ∼1.3 mIU/L at <40 years to ∼1.7 mIU/L at >70 years of age when we replaced TSH values among treated individuals. After exclusion of individuals with thyroid medication or disease, we observed a modest increase in geometric mean TSH in women from <40 years to >70 years of age (Figure 1B, Supplementary Table 2). The associations remained similar when we excluded the 761 (1.0%) TSH measurements in women that were performed during pregnancy (Supplementary Figure 1) and the 581 (0.4%) TSH measurements in women or men performed after any dispensing of amiodarone (Supplementary Figure 2), and when we excluded TSH measurements among current daily smokers (Supplementary Figure 3).

**Figure 1.**
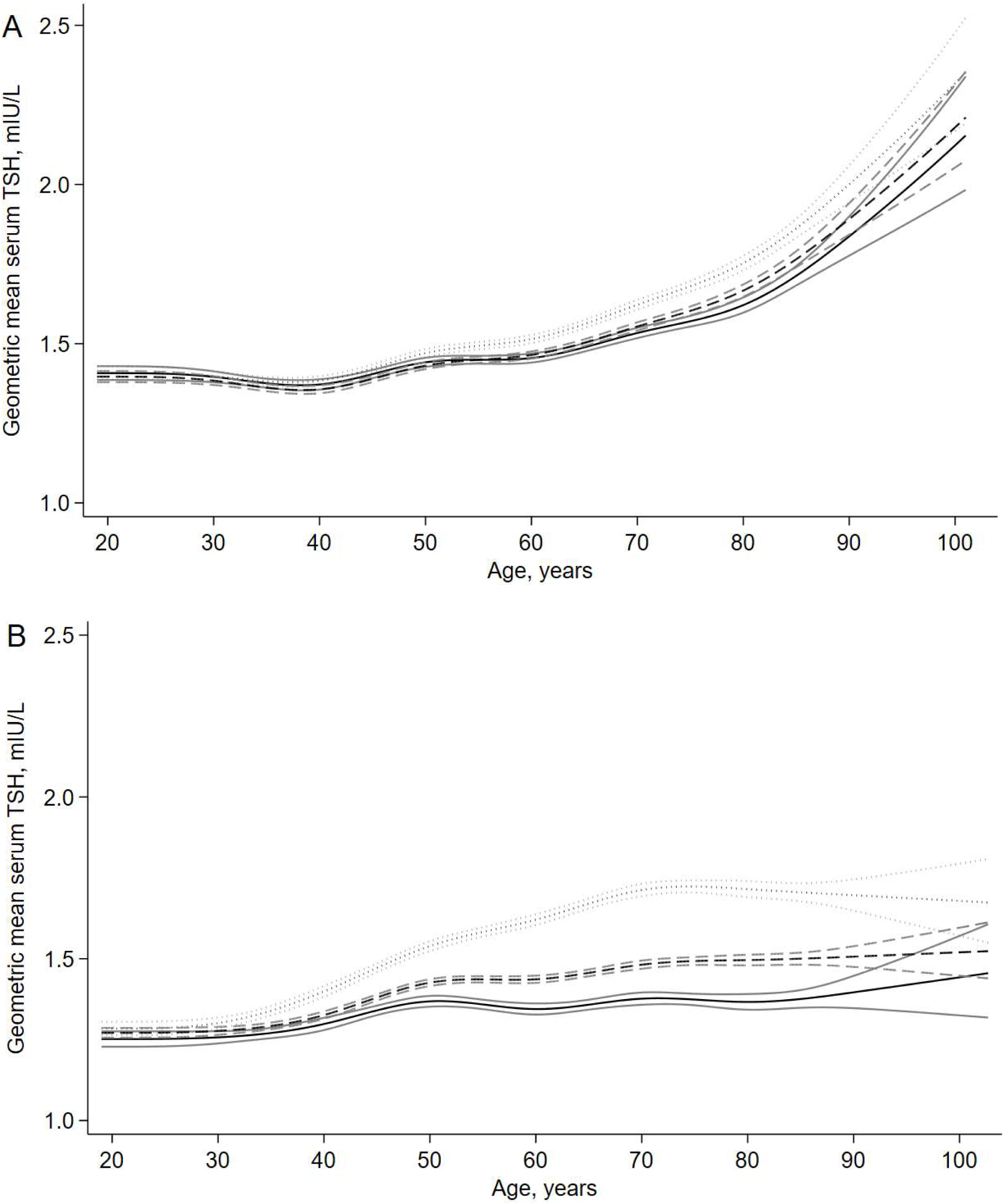
Estimated geometric mean serum TSH (with 95% confidence interval) by age, in men (A) and women (B). Results are presented for all TSH measurements (solid line), all TSH measurements with replacement of TSH values among people with thyroid medication (dotted line), and after exclusion of individuals with thyroid medication or disease (dashed line).

From 20 to 90 years of age, median (50-percentile) TSH increased by 0.6 mIU/L in men (Figure 2, Supplementary Table 3) and 0.4 mIU/L in women (Figure 2, Supplementary Table 4). There were signs of a widening TSH distribution by age in both men and women. Thus, among individuals without thyroid medication or disease, the 90^th^ percentile increased by 1.3 mIU/L in men and 0.9 mIU/L in women from 20 to 90 years of age.

**Figure 2.**
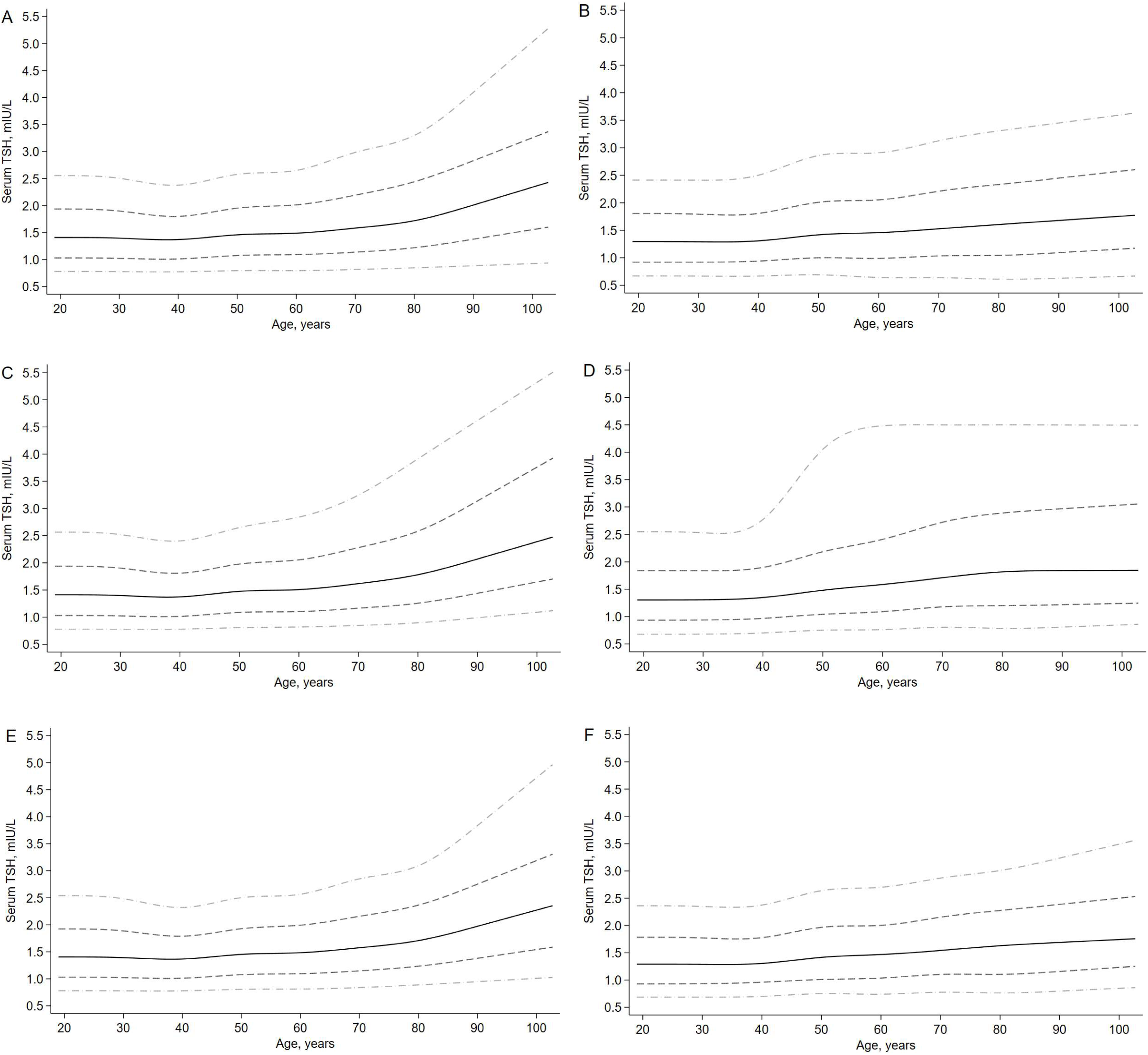
The estimated 10-, 25-, 50-, 75- and 90-percentiles of the serum TSH distribution by age in the overall study population of men (A, C and E) and women (B, D and F). Results are presented for all TSH measurements (A and B), all TSH measurements with replacement of TSH values among people with thyroid medication (C and D), and after exclusion of individuals with thyroid medication or disease (E and F).

In the strictly longitudinal analyses of within-individual changes in serum TSH, we observed that in men, the overall increase in geometric mean TSH from baseline was 2% at 11- year follow-up and 4% at 22-year follow-up, corresponding to TSH increases of 0.03 mIU/L and 0.06 mIU/L, respectively. In men without thyroid medication or disease, the analogous increases were 5% and 9%, corresponding to TSH increases of 0.07 mIU/L and 0.13 mIU/L, respectively. These changes were broadly similar across baseline ages <60 years, but older men had a stronger TSH increase between the 11-year and 22-year follow-up. In men aged ≥70 years at baseline, the increase in geometric mean TSH from baseline to the 22-year follow-up (when they were in their 90s) was 32% (GMR 1.32; 95% CI 1.18, 1.48; among men without thyroid medication or disease), corresponding to a TSH increase of 0.5 mIU/L (Table 2).

**Table 2.**
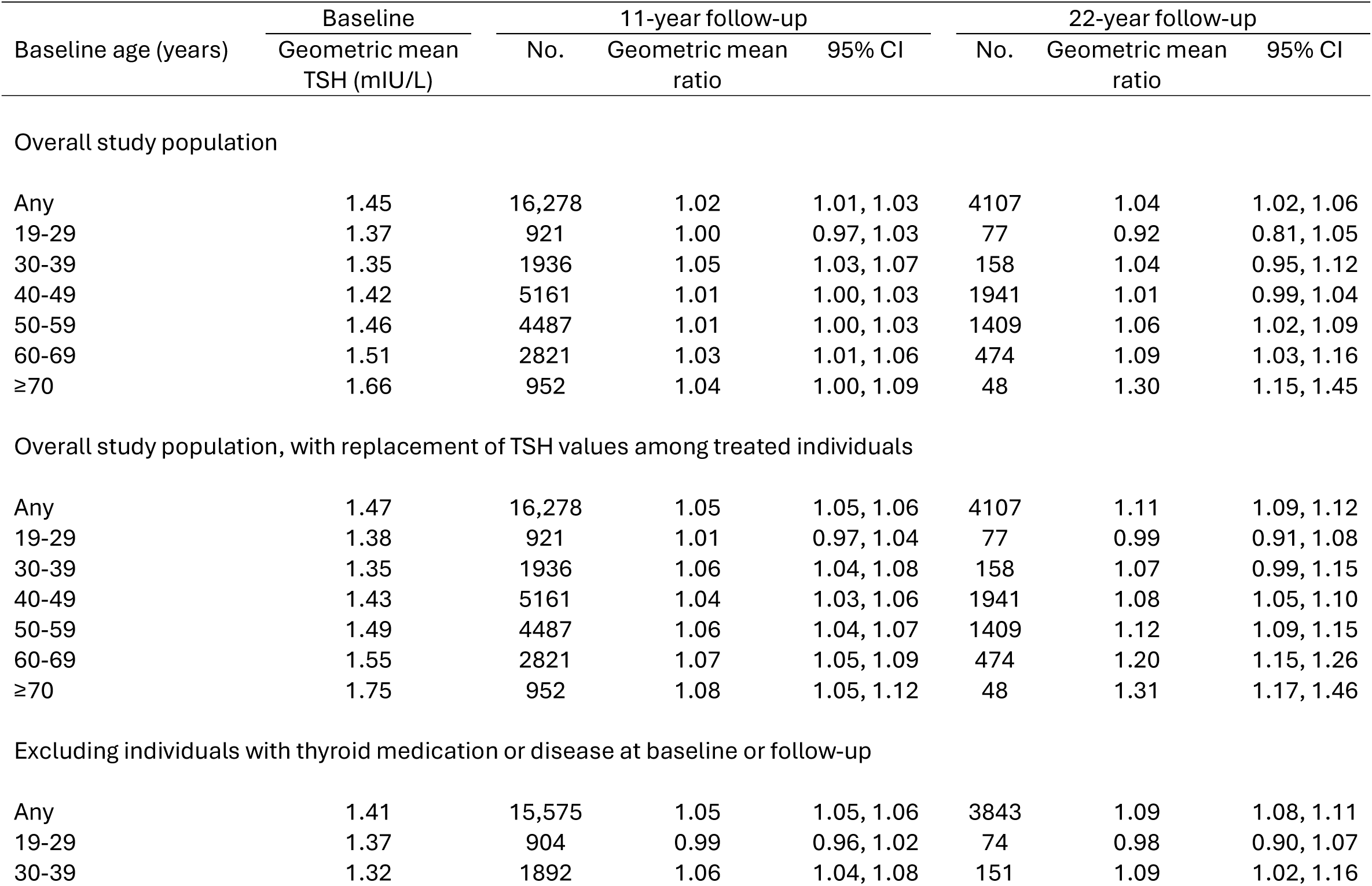
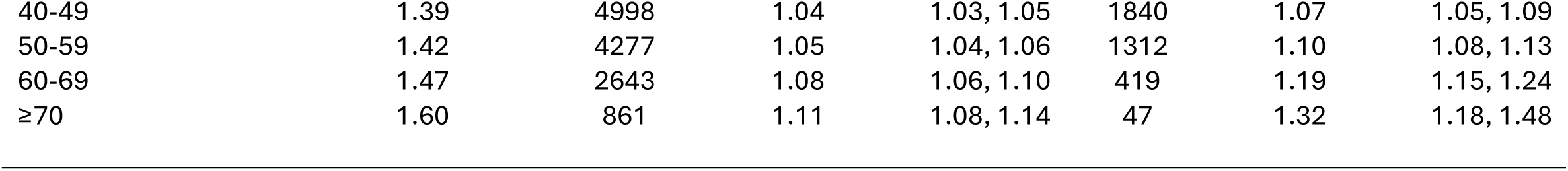
Within-individual changes in serum TSH from baseline (first HUNT survey with TSH measurement) to 11-year and 22-year follow-up, in 16,837 men with at least two TSH measurements. Changes in serum TSH are expressed by geometric mean ratios, which express the fold change in geometric mean serum TSH from baseline to follow-up.

| Baseline age (years) | Baseline | 11-year follow-up |  |  | 22-year follow-up |  |  |
| --- | --- | --- | --- | --- | --- | --- | --- |
|  | Geometric mean TSH (mIU/L) | No. | Geometric mean ratio | 95% CI | No. | Geometric mean ratio | 95% CI |
| Overall study population |  |  |  |  |  |  |  |
| Any | 1.45 | 16,278 | 1.02 | 1.01, 1.03 | 4107 | 1.04 | 1.02, 1.06 |
| 19-29 | 1.37 | 921 | 1.00 | 0.97, 1.03 | 77 | 0.92 | 0.81, 1.05 |
| 30-39 | 1.35 | 1936 | 1.05 | 1.03, 1.07 | 158 | 1.04 | 0.95, 1.12 |
| 40-49 | 1.42 | 5161 | 1.01 | 1.00, 1.03 | 1941 | 1.01 | 0.99, 1.04 |
| 50-59 | 1.46 | 4487 | 1.01 | 1.00, 1.03 | 1409 | 1.06 | 1.02, 1.09 |
| 60-69 | 1.51 | 2821 | 1.03 | 1.01, 1.06 | 474 | 1.09 | 1.03, 1.16 |
| ≥70 | 1.66 | 952 | 1.04 | 1.00, 1.09 | 48 | 1.30 | 1.15, 1.45 |
| Overall study population, with replacement of TSH values among treated individuals |  |  |  |  |  |  |  |
| Any | 1.47 | 16,278 | 1.05 | 1.05, 1.06 | 4107 | 1.11 | 1.09, 1.12 |
| 19-29 | 1.38 | 921 | 1.01 | 0.97, 1.04 | 77 | 0.99 | 0.91, 1.08 |
| 30-39 | 1.35 | 1936 | 1.06 | 1.04, 1.08 | 158 | 1.07 | 0.99, 1.15 |
| 40-49 | 1.43 | 5161 | 1.04 | 1.03, 1.06 | 1941 | 1.08 | 1.05, 1.10 |
| 50-59 | 1.49 | 4487 | 1.06 | 1.04, 1.07 | 1409 | 1.12 | 1.09, 1.15 |
| 60-69 | 1.55 | 2821 | 1.07 | 1.05, 1.09 | 474 | 1.20 | 1.15, 1.26 |
| ≥70 | 1.75 | 952 | 1.08 | 1.05, 1.12 | 48 | 1.31 | 1.17, 1.46 |
| Excluding individuals with thyroid medication or disease at baseline or follow-up |  |  |  |  |  |  |  |
| Any | 1.41 | 15,575 | 1.05 | 1.05, 1.06 | 3843 | 1.09 | 1.08, 1.11 |
| 19-29 | 1.37 | 904 | 0.99 | 0.96, 1.02 | 74 | 0.98 | 0.90, 1.07 |
| 30-39 | 1.32 | 1892 | 1.06 | 1.04, 1.08 | 151 | 1.09 | 1.02, 1.16 |
| 40-49 | 1.39 | 4998 | 1.04 | 1.03, 1.05 | 1840 | 1.07 | 1.05, 1.09 |
| 50-59 | 1.42 | 4277 | 1.05 | 1.04, 1.06 | 1312 | 1.10 | 1.08, 1.13 |
| 60-69 | 1.47 | 2643 | 1.08 | 1.06, 1.10 | 419 | 1.19 | 1.15, 1.24 |
| ≥70 | 1.60 | 861 | 1.11 | 1.08, 1.14 | 47 | 1.32 | 1.18, 1.48 |

In women, there was overall no within-individual increase in mean TSH. Conversely, there was an overall decline in geometric mean TSH of 7% from baseline to 11-year follow-up (GMR 0.93; 95% CI 0.93, 0.94) and 14% from baseline to 22-year follow-up (GMR 0.86; 95% CI 0.84, 0.88), corresponding to TSH declines of ∼0.10 mIU/L and ∼0.20 mIU/L, respectively. In women without thyroid medication or disease, geometric mean serum TSH remained virtually unchanged during follow-up. Only in the analyses where we replaced TSH values among treated individuals did we observe modest signs of within-individual increases in mean TSH during follow-up in women (Table 3). The within-individual TSH changes remained similar when we excluded current daily smokers, amiodarone users and pregnant women from the analysis (Supplementary Table 5).

**Table 3.** Within-individual changes in serum TSH from baseline (first HUNT survey with TSH measurement) to 11-year and 22-year follow-up, in 23,778 women with at least two TSH measurements. Changes in serum TSH are expressed by geometric mean ratios, which express the fold change in geometric mean serum TSH from baseline to follow-up.

| Baseline age (years) | Baseline | 11-year follow-up |  |  | 22-year follow-up |  |  |
| --- | --- | --- | --- | --- | --- | --- | --- |
|  | Geometric mean TSH (mIU/L) | No. | Geometric mean ratio | 95% CI | No. | Geometric mean ratio | 95% CI |
| Overall study population |  |  |  |  |  |  |  |
| Any | 1.39 | 22,692 | 0.93 | 0.93, 0.94 | 9506 | 0.86 | 0.84, 0.88 |
| 19-29 | 1.24 | 1436 | 0.97 | 0.93, 1.01 | 122 | 0.95 | 0.80, 1.11 |
| 30-39 | 1.23 | 3030 | 1.04 | 1.01, 1.07 | 218 | 0.92 | 0.79, 1.07 |
| 40-49 | 1.37 | 8866 | 0.94 | 0.92, 0.96 | 4513 | 0.88 | 0.85, 0.90 |
| 50-59 | 1.44 | 5339 | 0.90 | 0.88, 0.93 | 3229 | 0.85 | 0.82, 0.88 |
| 60-69 | 1.54 | 2882 | 0.86 | 0.83, 0.90 | 1241 | 0.81 | 0.77, 0.86 |
| ≥70 | 1.62 | 1139 | 0.88 | 0.84, 0.93 | 183 | 0.88 | 0.75, 1.04 |
| Overall study population, with replacement of TSH values among treated individuals |  |  |  |  |  |  |  |
| Any | 1.53 | 22,692 | 1.03 | 1.02, 1.04 | 9506 | 1.03 | 1.02, 1.05 |
| 19-29 | 1.27 | 1436 | 1.02 | 0.98, 1.06 | 122 | 1.09 | 0.95, 1.23 |
| 30-39 | 1.31 | 3030 | 1.10 | 1.07, 1.12 | 218 | 1.18 | 1.07, 1.31 |
| 40-49 | 1.48 | 8866 | 1.04 | 1.03, 1.06 | 4513 | 1.08 | 1.06, 1.10 |
| 50-59 | 1.64 | 5339 | 1.01 | 1.00, 1.03 | 3229 | 1.01 | 0.98, 1.03 |
| 60-69 | 1.80 | 2882 | 0.98 | 0.95, 1.00 | 1241 | 0.92 | 0.88, 0.96 |
| ≥70 | 1.87 | 1139 | 0.97 | 0.94, 1.00 | 183 | 0.93 | 0.83, 1.05 |
| Excluding individuals with thyroid medication or disease at baseline or follow-up |  |  |  |  |  |  |  |
| Any | 1.38 | 19,710 | 1.01 | 1.01, 1.02 | 7839 | 0.99 | 0.98, 1.00 |
| 19-29 | 1.24 | 1351 | 1.00 | 0.97, 1.03 | 108 | 1.00 | 0.90, 1.11 |
| 30-39 | 1.24 | 2782 | 1.08 | 1.06, 1.10 | 192 | 1.10 | 1.02, 1.18 |
| 40-49 | 1.36 | 7800 | 1.02 | 1.01, 1.03 | 3754 | 1.01 | 1.00, 1.03 |
| 50-59 | 1.45 | 4491 | 0.99 | 0.98, 1.00 | 2641 | 0.97 | 0.96, 0.99 |
| 60-69 | 1.52 | 2337 | 0.98 | 0.97, 1.00 | 995 | 0.95 | 0.92, 0.98 |
| ≥70 | 1.64 | 949 | 0.97 | 0.94, 1.00 | 149 | 0.93 | 0.85, 1.00 |

In both men and women without thyroid medication or disease at baseline or follow-up, the absolute deviations from the mean TSH change during follow-up increased by age (p<0.0001), indicating a widening of the distribution of increase in serum TSH at older age (Table 4).

**Table 4.** The mean (SD) increase in serum TSH from baseline (first HUNT survey with TSH measurement) to 11-year and 22-year follow-up, among men and women without thyroid medication or disease at baseline or follow-up, by age.

| Baseline age<br>(years) | TSH increase during<br>11-year follow-up (mIU/L) |  | TSH increase during<br>22-year follow-up (mIU/L) |  |
| --- | --- | --- | --- | --- |
|  | Mean<sup>a</sup> | SD<sup>b</sup> | Mean<sup>a</sup> | SD<sup>b</sup> |
| **Men** |  |  |  |  |
| Any | 0.09 | 0.63 | 0.18 | 0.69 |
| 19-29 | -0.01 | 0.69 | -0.02 | 0.63 |
| 30-39 | 0.09 | 0.58 | 0.13 | 0.69 |
| 40-49 | 0.07 | 0.59 | 0.12 | 0.65 |
| 50-59 | 0.09 | 0.62 | 0.20 | 0.68 |
| 60-69 | 0.15 | 0.67 | 0.36 | 0.80 |
| ≥70 | 0.21 | 0.73 | 0.63 | 0.93 |
| *p for trend* | <0.0001 | <0.0001 | <0.0001 | <0.0001 |
| **Women** |  |  |  |  |
| Any | 0.03 | 0.67 | 0.03 | 0.74 |
| 19-29 | 0.00 | 0.70 | 0.01 | 0.79 |
| 30-39 | 0.12 | 0.64 | 0.13 | 0.74 |
| 40-49 | 0.03 | 0.64 | 0.06 | 0.70 |
| 50-59 | 0.00 | 0.68 | 0.00 | 0.74 |
| 60-69 | -0.01 | 0.69 | -0.01 | 0.85 |
| ≥70 | -0.03 | 0.75 | -0.03 | 0.89 |
| *p for trend* | <0.0001 | 0.0001 | 0.001 | <0.0001 |

SD, standard deviation; TSH, thyroid-stimulating hormone
<sup>a</sup> The p-value is for trend in mean serum TSH change across age categories
<sup>b</sup> The p-value is for trend in absolute deviations from the mean TSH change across age categories

## Discussion

In this study, we examined age-related changes in serum TSH concentrations during long-term follow-up of a population-based cohort. In analyses integrating all cross-sectional and longitudinal data, mean TSH was higher at older age in men and, weaker and less consistently, in women. The TSH distribution was wider at older age in both men and women. The mean within-individual increase in TSH during 22-year follow-up was modest at 0.13 mIU/L in men without thyroid medication or disease. At ages >70 years this increase was stronger, with men in their 70s at baseline having a TSH increase of 0.5 mIU/L until their 90s. In women, there were no consistent signs of within-individual increases in mean TSH by age, and overall mean TSH changed by only 0.01 mIU/L from baseline to the 22-year follow-up in women without thyroid medication or disease.

In the Australian Busselton Health Study, mean serum TSH increased by 0.32 mIU/L during 13-year follow-up of 908 adults without thyroid disease or antibodies, and this increase was 0.08 mIU/L stronger for each decade of age.^7^ Among 657 elderly individuals in the Cardiovascular Health Study All Stars Study in the USA, mean serum TSH increased by 0.34 mIU/L during 13-year follow-up until a mean age of 85 years.^8^ Among 204 participants of the English Whickham cohort study with a mean baseline age of 77 years, mean TSH increased by 0.29 mIU/L during 8-year follow-up, and this increase was 0.02 mIU/L stronger per one-year older age at baseline.^9^ In other longitudinal studies with 5-7 years of follow-up, changes in TSH were not detected. ^10–12^ Our estimated TSH increase in elderly men between their 70s and 90s is compatible to the longitudinal change observed in the elderly in the Cardiovascular Health Study All Stars Study and the Whickham cohort study. However, our overall TSH increase during 22-year follow-up in the general adult population is smaller than that observed during 13 years in the Busselton Health Study.

A strength of this study is that we were able to estimate within-individual TSH changes during long-term follow-up in a population-based sample, extending individual-level follow-up to more than 20 years. Our large sample size enabled precise estimates both overall and across age groups. Utilizing method comparisons of the TSH measurement methods across the surveys, we could minimize the impact of any systematic differences in measurement methods during the follow-up period. Nevertheless, we cannot exclude that minor systematic differences between the measurement methods may have influenced the results. Also, as free thyroxine and TPO antibody measurements were not available in euthyroid individuals, we cannot exclude that occult early-stage thyroid disease may have contributed to the estimated changes in TSH. However, we would assume that such influence of occult thyroid disease would be present primarily in women, in whom autoimmune thyroid disease is much more common. In contrast, we observed the strongest and most consistent age-TSH associations in men. We could not account for the impact on serum TSH from severe non-thyroidal illness or medication other than thyroid medication and amiodarone. Such factors, occurring more commonly at older age, may have contributed to the changes in distribution of TSH levels at older age. The TSH distribution may depend on past and current iodine status, and our results may not be generalizable to countries with other history of iodine sufficiency.

The sex difference with a weaker and less consistent age-TSH association in women could be influenced by more widespread use of thyroid hormone replacement therapy in women during follow-up. We have previously observed that the age-adjusted prevalence of thyroid hormone replacement therapy increased between the HUNT2 and HUNT3 surveys, more so in women (3 percentage points) than in men (1 percentage point), accompanied by a similar decline in the prevalence of mildly elevated TSH concentrations.^18^ We have also shown that already at HUNT2, individuals with a genetic predisposition to higher physiological serum TSH concentrations were more likely to receive thyroid hormone replacement therapy.^21^ Thus, it is possible that more widespread use of thyroid hormone replacement therapy may have made the population relatively replete of individuals with higher physiological serum TSH, which may have attenuated the estimates of the age-TSH association. Consistent with that possibility, we observed signs of higher TSH at older age in women when we replaced their TSH values to approximate the TSH level they presumably had prior to treatment.

Although the mean within-individual increase in TSH during follow-up was relatively modest in our study, we observed wider TSH distributions at older age when integrating all our cross-sectional and longitudinal TSH measurements. Thus, among individuals without thyroid disease or medication, the 90^th^ percentile increased substantially from 20 to 90 years of age, by 1.3 mIU/L in men and 0.9 mIU/L in women. Our observations corroborate previous cross-sectional^2,5,22^ and longitudinal^9^ studies suggesting that higher upper reference limits for TSH may be warranted at older age.

In conclusion, 22-year follow-up of our population-based cohort showed a mean within-individual TSH increase in men that was modest overall, but stronger in men followed from their 70s to their 90s. In women, there was no consistent evidence that mean serum TSH increased during follow-up, but more frequent thyroid hormone supplementation may have skewed the TSH distribution away from higher, yet physiological levels. The TSH distribution was wider at older age in both men and women, supporting the suggestion that age-specific reference ranges for serum TSH may be warranted.

## Acknowledgements

The Trøndelag Health Study (HUNT) is a collaboration between HUNT Research Centre (Faculty of Medicine and Health Sciences, NTNU, Norwegian University of Science and Technology), Trøndelag County Council, Central Norway Regional Health Authority, and the Norwegian Institute of Public Health.

## Data Availability Statement

Due to privacy protection for participants, individual-level data from the HUNT Study are not publicly available. For information on how to apply for data from the HUNT Study, please see: https://www.ntnu.edu/hunt/data.

