## Supplementary Figure 1 for "Changes in serum TSH across the adult lifespan: 22-year follow-up of the HUNT Study"

**Supplementary Figure 1.** Estimated geometric mean serum TSH (with 95% confidence interval) by age in women, including all TSH measurments (solid line) and excluding TSH measurements performed during pregnancy (dashed line).

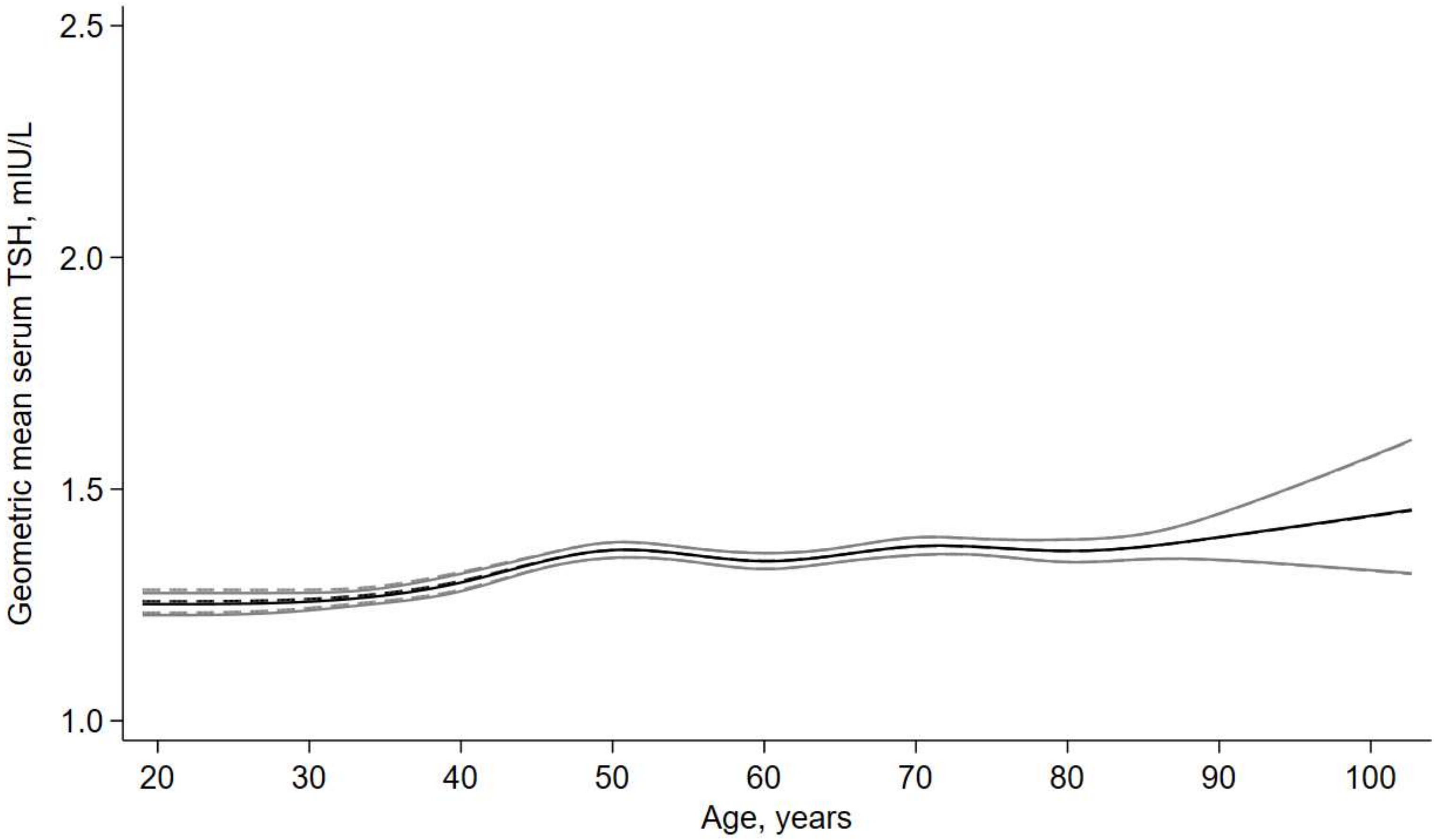
