## Supplementary Figure 2 for "Changes in serum TSH across the adult lifespan: 22-year follow-up of the HUNT Study"

**Supplementary Figure 2.** Estimated geometric mean serum TSH (with 95% confidence interval) by age, in men (A) and women (B), including all TSH measurments (solid line) and excluding TSH measurements performed after any dispensing of amiodarone between 2004 and the date of TSH measurement (dashed line).

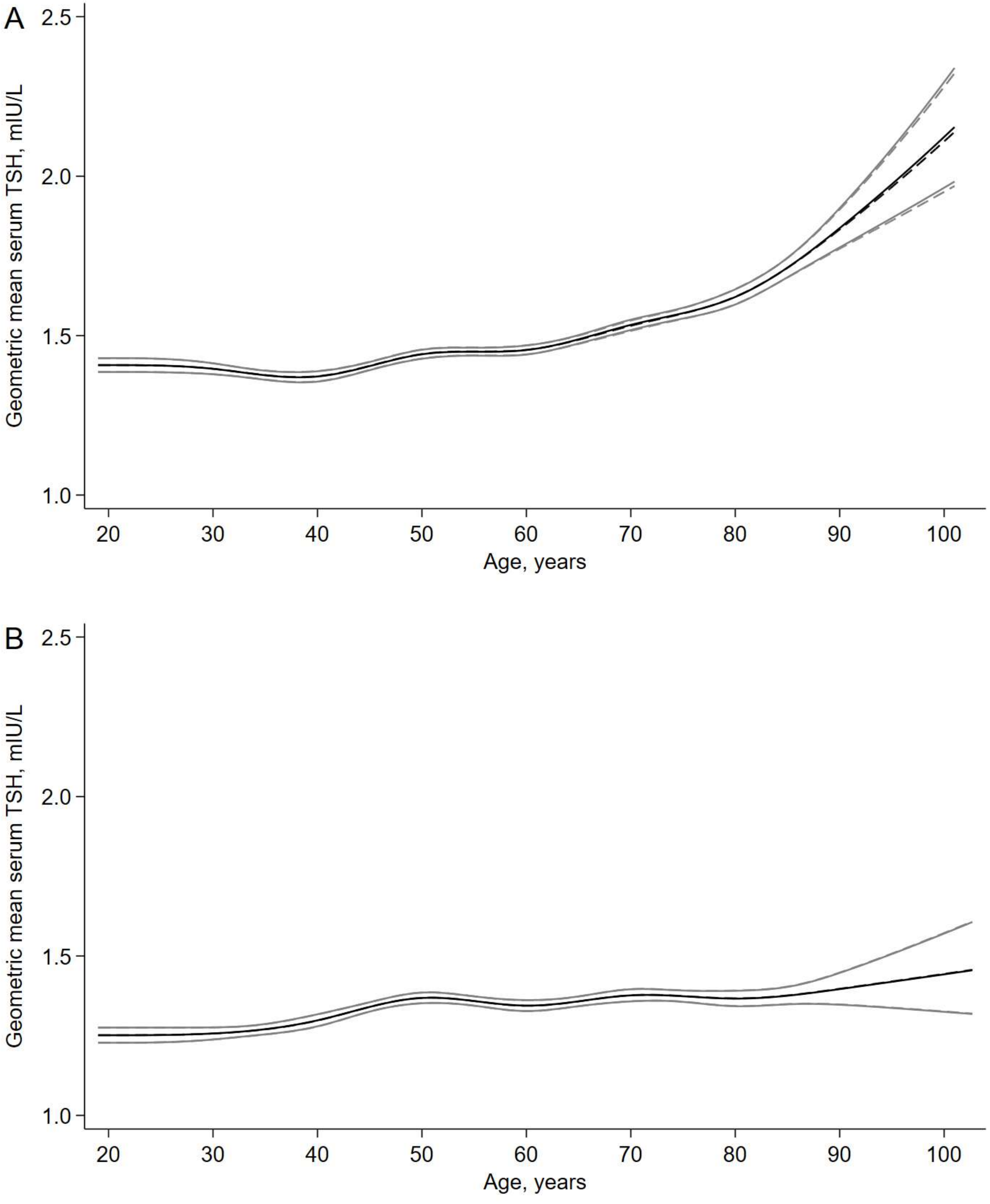
