## Supplementary Figure 3 for "Changes in serum TSH across the adult lifespan: 22-year follow-up of the HUNT Study"

**Supplementary Figure 3.** Estimated geometric mean serum TSH (with 95% confidence interval) by age, in men (A) and women (B), including all TSH measurments (solid line) and excluding TSH measurements among current daily smokers (dashed line).

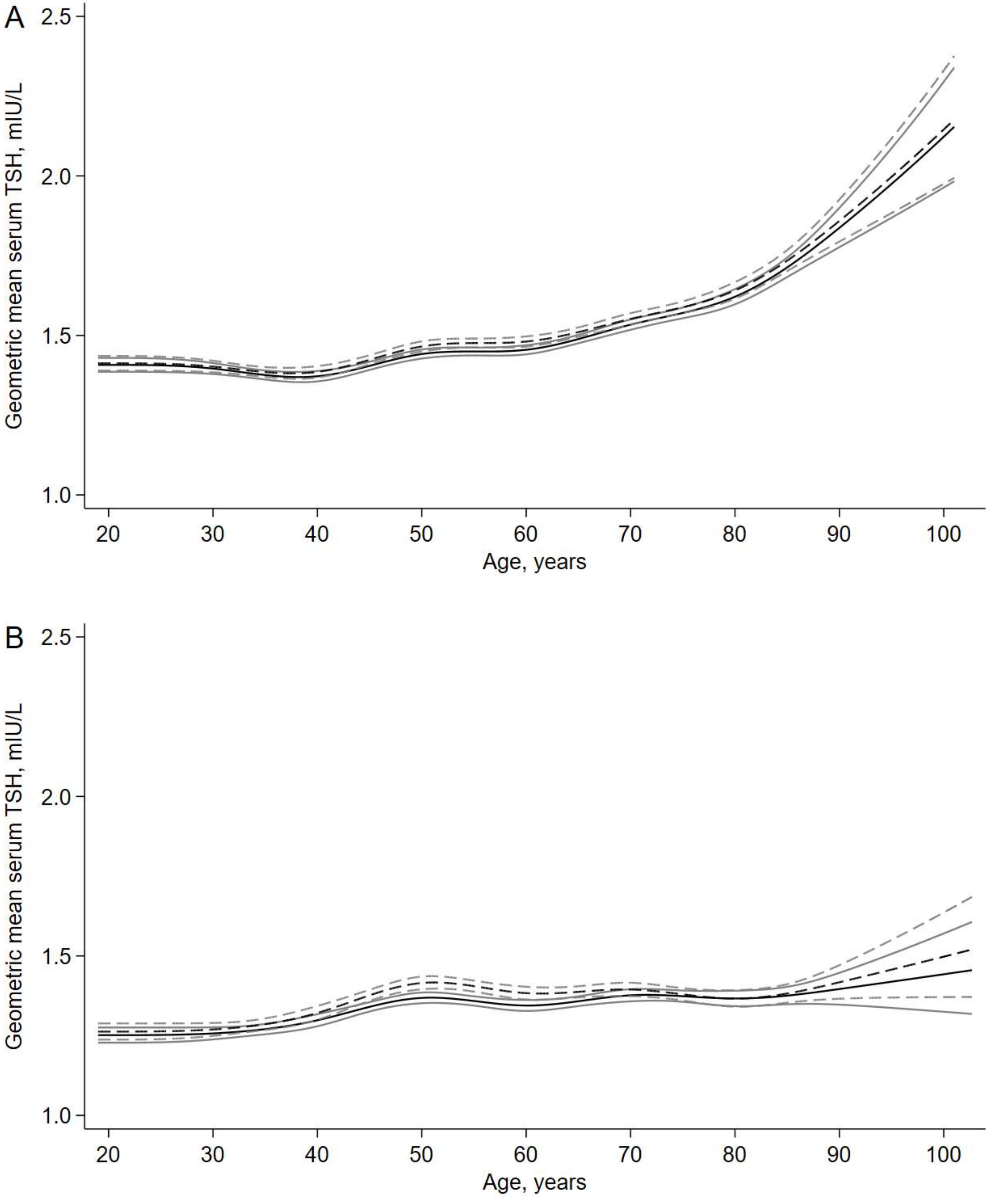
