## Supplementary Table 1 for "Changes in serum TSH across the adult lifespan: 22-year follow-up of the HUNT Study"

**Supplementary Table 1.** Number of individuals with TSH measurement, by age group, sex and survey.

| Age, years | HUNT2 (1995-97) | | HUNT3 (2006-08) | | HUNT4 (2017-19) | |
|---|---|---|---|---|---|---|
| | Men | Women | Men | Women | Men | Women |
| 19-29[a] | 192 | 229 | 1763 | 2472 | 2554 | 3354 |
| 30-39 | 263 | 335 | 2765 | 3881 | 2711 | 3715 |
| 40-49 | 3007 | 6571 | 4415 | 5283 | 3721 | 4913 |
| 50-59 | 2659 | 5627 | 5271 | 5831 | 4748 | 5656 |
| 60-69 | 2201 | 4630 | 4561 | 4978 | 5301 | 5644 |
| 70-79 | 1782 | 4265 | 2612 | 2996 | 3874 | 4242 |
| 80-89 | 511 | 1522 | 920 | 1282 | 1272 | 1639 |
| ≥90 | 28 | 92 | 47 | 98 | 168 | 324 |

[a] All residents aged ≥20 years at the estimated time of survey participation were invited, and some were aged 19 years at the date of participation
