## Supplementary Table 2 for "Changes in serum TSH across the adult lifespan: 22-year follow-up of the HUNT Study"

**Supplementary Table 2.** Estimated geometric mean serum TSH (with 95% confidence interval) at specific ages, in men and women.

| Age (years) | Men | | | Women | | |
|---|---|---|---|---|---|---|
| | Geometric mean TSH (mIU/L) | 95% CI | | Geometric mean TSH (mIU/L) | 95% CI | |
| Overall study population | | | | | | |
| 20 | 1.41 | 1.39 | 1.43 | 1.25 | 1.23 | 1.28 |
| 30 | 1.40 | 1.38 | 1.41 | 1.26 | 1.24 | 1.28 |
| 40 | 1.37 | 1.36 | 1.39 | 1.30 | 1.28 | 1.32 |
| 50 | 1.44 | 1.43 | 1.46 | 1.37 | 1.35 | 1.39 |
| 60 | 1.45 | 1.44 | 1.47 | 1.34 | 1.33 | 1.36 |
| 70 | 1.53 | 1.52 | 1.55 | 1.38 | 1.36 | 1.40 |
| 80 | 1.62 | 1.60 | 1.65 | 1.37 | 1.34 | 1.39 |
| 90 | 1.84 | 1.78 | 1.90 | 1.40 | 1.35 | 1.45 |
| 100 | 2.12 | 1.96 | 2.30 | 1.44 | 1.32 | 1.58 |
| Overall study population, with replacement of TSH values among treated individuals | | | | | | |
| 20 | 1.41 | 1.39 | 1.43 | 1.28 | 1.26 | 1.31 |
| 30 | 1.40 | 1.38 | 1.41 | 1.30 | 1.28 | 1.32 |
| 40 | 1.38 | 1.37 | 1.40 | 1.40 | 1.38 | 1.42 |
| 50 | 1.47 | 1.46 | 1.48 | 1.54 | 1.52 | 1.55 |
| 60 | 1.51 | 1.50 | 1.53 | 1.62 | 1.60 | 1.64 |
| 70 | 1.62 | 1.61 | 1.64 | 1.71 | 1.69 | 1.73 |
| 80 | 1.75 | 1.73 | 1.78 | 1.72 | 1.69 | 1.74 |
| 90 | 2.00 | 1.94 | 2.06 | 1.70 | 1.65 | 1.75 |
| 100 | 2.32 | 2.17 | 2.48 | 1.68 | 1.57 | 1.79 |
| Overall study population after exclusion of measurements in individuals with thyroid medication or disease | | | | | | |
| 20 | 1.40 | 1.38 | 1.41 | 1.27 | 1.26 | 1.29 |
| 30 | 1.38 | 1.37 | 1.40 | 1.28 | 1.26 | 1.29 |
| 40 | 1.36 | 1.34 | 1.37 | 1.33 | 1.31 | 1.34 |
| 50 | 1.43 | 1.42 | 1.44 | 1.43 | 1.42 | 1.44 |
| 60 | 1.46 | 1.45 | 1.48 | 1.44 | 1.43 | 1.45 |
| 70 | 1.55 | 1.54 | 1.57 | 1.48 | 1.47 | 1.49 |
| 80 | 1.67 | 1.65 | 1.69 | 1.50 | 1.48 | 1.51 |
| 90 | 1.89 | 1.84 | 1.94 | 1.51 | 1.47 | 1.54 |
| 100 | 2.18 | 2.05 | 2.31 | 1.52 | 1.45 | 1.60 |
