## Supplementary Table 3 for "Changes in serum TSH across the adult lifespan: 22-year follow-up of the HUNT Study"

**Supplementary Table 3.** Estimated percentiles of the serum TSH distribution (in mIU/L) at specific ages, in men.

| Age (years) | Percentiles | | | | |
|---|---|---|---|---|---|
| | 10 | 25 | 50 | 75 | 90 |
| Overall study population | | | | | |
| 20 | 0.78 | 1.03 | 1.41 | 1.94 | 2.55 |
| 30 | 0.78 | 1.02 | 1.40 | 1.90 | 2.51 |
| 40 | 0.77 | 1.01 | 1.37 | 1.80 | 2.38 |
| 50 | 0.80 | 1.08 | 1.46 | 1.95 | 2.58 |
| 60 | 0.80 | 1.09 | 1.49 | 2.01 | 2.65 |
| 70 | 0.82 | 1.14 | 1.58 | 2.19 | 2.99 |
| 80 | 0.85 | 1.22 | 1.72 | 2.44 | 3.30 |
| 90 | 0.89 | 1.38 | 2.01 | 2.83 | 4.10 |
| 100 | 0.93 | 1.55 | 2.34 | 3.26 | 5.03 |
| Overall study population, with replacement of TSH values among treated individuals | | | | | |
| 20 | 0.78 | 1.03 | 1.41 | 1.94 | 2.57 |
| 30 | 0.78 | 1.02 | 1.40 | 1.90 | 2.52 |
| 40 | 0.78 | 1.01 | 1.37 | 1.81 | 2.40 |
| 50 | 0.81 | 1.09 | 1.48 | 1.98 | 2.65 |
| 60 | 0.82 | 1.10 | 1.51 | 2.06 | 2.84 |
| 70 | 0.85 | 1.17 | 1.62 | 2.28 | 3.25 |
| 80 | 0.90 | 1.26 | 1.78 | 2.58 | 3.91 |
| 90 | 0.99 | 1.44 | 2.07 | 3.14 | 4.62 |
| 100 | 1.09 | 1.65 | 2.39 | 3.76 | 5.32 |
| After exclusion of measurements in individuals with thyroid medication or disease | | | | | |
| 20 | 0.78 | 1.03 | 1.40 | 1.92 | 2.54 |
| 30 | 0.78 | 1.02 | 1.39 | 1.89 | 2.48 |
| 40 | 0.78 | 1.01 | 1.37 | 1.79 | 2.32 |
| 50 | 0.81 | 1.08 | 1.45 | 1.93 | 2.50 |
| 60 | 0.81 | 1.10 | 1.48 | 1.99 | 2.57 |
| 70 | 0.84 | 1.15 | 1.58 | 2.16 | 2.85 |
| 80 | 0.89 | 1.23 | 1.71 | 2.37 | 3.09 |
| 90 | 0.95 | 1.38 | 1.97 | 2.75 | 3.84 |
| 100 | 1.01 | 1.54 | 2.27 | 3.19 | 4.72 |
