## Supplementary Table 4 for "Changes in serum TSH across the adult lifespan: 22-year follow-up of the HUNT Study"

**Supplementary Table 4.** Estimated percentiles of the serum TSH distribution (in mIU/L) at specific ages, in women.

| Age (years) | Percentiles | | | | |
|---|---|---|---|---|---|
| | 10 | 25 | 50 | 75 | 90 |
| Overall study population | | | | | |
| 20 | 0.67 | 0.92 | 1.29 | 1.80 | 2.41 |
| 30 | 0.67 | 0.92 | 1.29 | 1.79 | 2.41 |
| 40 | 0.67 | 0.94 | 1.31 | 1.81 | 2.50 |
| 50 | 0.69 | 1.00 | 1.42 | 2.01 | 2.86 |
| 60 | 0.64 | 0.99 | 1.46 | 2.05 | 2.91 |
| 70 | 0.64 | 1.03 | 1.53 | 2.21 | 3.13 |
| 80 | 0.61 | 1.04 | 1.60 | 2.33 | 3.31 |
| 90 | 0.63 | 1.09 | 1.68 | 2.45 | 3.45 |
| 100 | 0.66 | 1.16 | 1.75 | 2.57 | 3.59 |
| Overall study population, with replacement of TSH values among treated individuals | | | | | |
| 20 | 0.68 | 0.94 | 1.30 | 1.84 | 2.55 |
| 30 | 0.68 | 0.94 | 1.31 | 1.84 | 2.53 |
| 40 | 0.70 | 0.97 | 1.35 | 1.90 | 2.77 |
| 50 | 0.75 | 1.04 | 1.48 | 2.18 | 4.05 |
| 60 | 0.76 | 1.09 | 1.59 | 2.41 | 4.48 |
| 70 | 0.81 | 1.18 | 1.71 | 2.72 | 4.50 |
| 80 | 0.79 | 1.20 | 1.82 | 2.89 | 4.50 |
| 90 | 0.81 | 1.22 | 1.84 | 2.97 | 4.50 |
| 100 | 0.85 | 1.24 | 1.84 | 3.04 | 4.49 |
| After exclusion of measurements in individuals with thyroid medication or disease | | | | | |
| 20 | 0.68 | 0.93 | 1.29 | 1.78 | 2.36 |
| 30 | 0.68 | 0.93 | 1.29 | 1.77 | 2.35 |
| 40 | 0.70 | 0.96 | 1.30 | 1.78 | 2.37 |
| 50 | 0.75 | 1.01 | 1.41 | 1.97 | 2.64 |
| 60 | 0.74 | 1.04 | 1.47 | 2.00 | 2.70 |
| 70 | 0.78 | 1.10 | 1.54 | 2.15 | 2.87 |
| 80 | 0.76 | 1.10 | 1.63 | 2.28 | 3.01 |
| 90 | 0.80 | 1.16 | 1.69 | 2.39 | 3.24 |
| 100 | 0.85 | 1.23 | 1.74 | 2.50 | 3.49 |
