## Supplementary Table 5 for "Changes in serum TSH across the adult lifespan: 22-year follow-up of the HUNT Study"

**Supplementary Table 5.** Within-individual changes in serum TSH from baseline (first HUNT survey with TSH measurement) to 11-year and 22-year follow-up, after exclusion of TSH measurements in current daily smokers or during pregnancy. Changes in serum TSH are expressed by geometric mean ratios, which express the fold change in geometric mean serum TSH from baseline to follow-up.

| | Baseline | 11-year follow-up | | | 22-year follow-up | | |
|---|---|---|---|---|---|---|---|
| Baseline age (years) | Geometric mean TSH (mIU/L) | No. | Geometric mean ratio | 95% CI | No. | Geometric mean ratio | 95% CI |
| Among men who were not current daily smokers at baseline or follow-up | | | | | | | |
| Any | 1.48 | 12,944 | 1.02 | 1.01, 1.03 | 3089 | 1.04 | 1.01, 1.06 |
| 19-29 | 1.36 | 790 | 1.00 | 0.96, 1.04 | 61 | 0.89 | 0.77, 1.04 |
| 30-39 | 1.36 | 1662 | 1.05 | 1.03, 1.08 | 125 | 1.05 | 0.97, 1.14 |
| 40-49 | 1.45 | 4018 | 1.02 | 1.00, 1.03 | 1384 | 1.02 | 0.99, 1.06 |
| 50-59 | 1.51 | 3438 | 1.01 | 0.99, 1.03 | 1080 | 1.04 | 1.00, 1.08 |
| 60-69 | 1.55 | 2241 | 1.03 | 1.00, 1.05 | 396 | 1.10 | 1.03, 1.18 |
| ≥70 | 1.67 | 795 | 1.05 | 1.00, 1.10 | 43 | 1.31 | 1.15, 1.48 |
| Among women who were not current daily smokers at baseline or follow-up | | | | | | | |
| Any | 1.45 | 16,464 | 0.93 | 0.92, 0.95 | 6765 | 0.84 | 0.82, 0.86 |
| 19-29 | 1.26 | 1165 | 0.98 | 0.93, 1.03 | 88 | 1.00 | 0.83, 1.21 |
| 30-39 | 1.25 | 2467 | 1.07 | 1.04, 1.10 | 147 | 0.89 | 0.73, 1.08 |
| 40-49 | 1.45 | 6050 | 0.95 | 0.93, 0.97 | 2951 | 0.87 | 0.84, 0.91 |
| 50-59 | 1.54 | 3701 | 0.88 | 0.85, 0.90 | 2394 | 0.80 | 0.77, 0.83 |
| 60-69 | 1.59 | 2135 | 0.86 | 0.82, 0.89 | 1018 | 0.81 | 0.76, 0.86 |
| ≥70 | 1.66 | 946 | 0.89 | 0.84, 0.94 | 167 | 0.89 | 0.75, 1.06 |
| Among women who were not pregnant at baseline or follow-up | | | | | | | |
| Any | 1.40 | 22,341 | 0.93 | 0.92, 0.94 | 9473 | 0.86 | 0.84, 0.88 |
| 19-29 | 1.26 | 1244 | 0.97 | 0.93, 1.01 | 111 | 0.93 | 0.77, 1.11 |
| 30-39 | 1.24 | 2903 | 1.04 | 1.01, 1.07 | 209 | 0.90 | 0.77, 1.06 |

| | | | | | | | |
|---|---|---|---|---|---|---|---|
| 40-49 | 1.37 | 8834 | 0.94 | 0.92, 0.96 | 4500 | 0.88 | 0.85, 0.90 |
| Among men with no dispensing of amiodarone between 2004 and follow-up | | | | | | | |
| Any | 1.45 | 16,174 | 1.02 | 1.01, 1.03 | 4027 | 1.04 | 1.02, 1.06 |
| 19-29 | 1.37 | 920 | 1.00 | 0.97, 1.03 | 77 | 0.92 | 0.81, 1.05 |
| 30-39 | 1.35 | 1936 | 1.05 | 1.03, 1.07 | 156 | 1.04 | 0.96, 1.13 |
| 40-49 | 1.42 | 5148 | 1.01 | 1.00, 1.03 | 1917 | 1.01 | 0.98, 1.04 |
| 50-59 | 1.46 | 4457 | 1.01 | 1.00, 1.03 | 1371 | 1.05 | 1.02, 1.09 |
| 60-69 | 1.51 | 2777 | 1.03 | 1.01, 1.05 | 458 | 1.10 | 1.04, 1.17 |
| ≥70 | 1.66 | 936 | 1.04 | 1.00, 1.09 | 48 | 1.30 | 1.15, 1.45 |
| Among women with no dispensing of amiodarone between 2004 and follow-up | | | | | | | |
| Any | 1.39 | 22,679 | 0.93 | 0.92, 0.94 | 9470 | 0.86 | 0.84, 0.88 |
| 19-29 | 1.24 | 1436 | 0.97 | 0.93, 1.01 | 122 | 0.95 | 0.80, 1.11 |
| 30-39 | 1.23 | 3029 | 1.04 | 1.02, 1.07 | 218 | 0.92 | 0.79, 1.07 |
| 40-49 | 1.37 | 8864 | 0.94 | 0.92, 0.96 | 4502 | 0.88 | 0.85, 0.90 |
| 50-59 | 1.44 | 5335 | 0.90 | 0.88, 0.93 | 3213 | 0.85 | 0.82, 0.88 |
| 60-69 | 1.53 | 2877 | 0.86 | 0.83, 0.90 | 1232 | 0.81 | 0.77, 0.86 |
| ≥70 | 1.61 | 1138 | 0.88 | 0.84, 0.93 | 183 | 0.88 | 0.75, 1.04 |
